# Assessment of magnitude and spectrum of cardiovascular disease admissions and outcomes in Saint Paul Hospital Millennium Medical College, Addis Ababa: A retrospective study

**DOI:** 10.1101/2022.04.12.22273778

**Authors:** Mekoya D. Mengistu, Henok Benti

## Abstract

**Background:** Cardiovascular diseases remain the leading cause of death in the world and approximately 80% of all cardiovascular-related deaths occur in low and middle income countries including Ethiopia.

**Methods:** The aim of the study was to assess the magnitude and spectrum of cardiovascular admissions and its outcomes among medical patients admitted to both Medical Ward and ICU of St. Paul Teaching Hospital from 1st of Jan 2020 to 1st of Jan 2021.

**Results:** Out of 1,165 annual medical admissions, the prevalence of cardiovascular diseases(CVD) was 30.3%. About 60%(212) of patients had advanced congestive heart failure of diverse causes. Hypertensive heart disease (HHD) was the next predominant diagnosis (41%), and also the leading cause of cardiac diseases followed by rheumatic valvular heart disease(RVHD) (18%) and Ischemic heart disease (IHD) (12.2%), respectively. Yong age, rural residence and female sex were associated with RVHD(p=0.001). Stroke also accounted for 20% of CVD admission (hemorrhagic stroke-17% Vs Ischemic stroke-83%). Hypertension was the predominate risk factor for CVD and present in 46.7%(168) of patients. The mean hospital stay was 12days and in hospital mortality rate was 24.3%, septic shock being the commonest immediate cause of death followed by fatal arrhythmia, brain herniation, and massive PTE.

**Conclusion:** Cardiovascular diseases were highly prevalent in the study area causing significant morbidity and mortality. Therefore, Comprehensive approach is needed to timely screen for risk reduction, delay or prevent diseases development and subsequent complications.

## Introduction

The adverse human, social and economic consequences of non-communicable diseases (NCDs) are felt by all societies and economies, but they are particularly devastating in poor and vulnerable populations [1]. NCDs currently cause more deaths than all other causes combined and NCD deaths are projected to increase from 38 million in 2012 to 52 million by 2030. Nearly three quarters of all NCD deaths occur in low- and middle-income countries. The four major NCDs responsible for 82% of NCD deaths in 2012 were cardiovascular diseases, cancers, respiratory diseases including asthma and chronic obstructive pulmonary disease, and diabetes [2]. These four diseases are the world’s biggest killers, causing an estimated 35 million deaths each year - 60% of all deaths globally - with 80% in low- and middle-income countries, which also have a high proportion of deaths in middle age [1]. NCD deaths will increase by 17% over the next ten years and the greatest increase will be seen in the African region[2]. In Ethiopia, the recent NCD deaths are estimated at around 42% [3]. WHO also estimated in 2011 that 34% of Ethiopian population is dying from non-communicable diseases, with a national cardiovascular disease prevalence of 15%[4, 5].

The epidemic of cardiovascular disease (CVD) is a global phenomenon, and the magnitude in terms of incidence and prevalence increases in the developing world. The cardiovascular disease remains the leading cause of death in the world and approximately 80% of all cardiovascular-related deaths occur in low and middle income countries and at a younger age in comparison to high-income countries [6]. Africa is home to for over 1 billion people, and is a major contributor to the global burden of CVD [7, 8]. In 2013, an estimated 1 million deaths were attributable to CVD in sub-Saharan Africa alone, which constituted 5.5% of all global CVD-related deaths and 11.3% of all deaths in Africa [7, 9]. CVD-related deaths contributed to 38% of all non-communicable disease-related deaths in Africa, reflecting the growing threat of both non-communicable disease and CVD. An almost twofold increase in the over-all number of CVD-related deaths since 1990 has been reported, with a >10% difference in mortality among women compared with men [9]

The cardiovascular spectrum study in Addis Ababa during 2014 pointed out the five most common cardiovascular diseases; valvular heart disease (62%), hypertension (14.7%), cerebrovascular disease (11.5%), congenital heart disease (8.5%), and ischemic heart disease (IHD) (6.8%) [10]. Of all the 300 cases of heart failure, 81.3% of the advanced heart failure (New York Heart Association (NYHA) class III or IV) patients had a primary diagnosis of valvular heart disease which predominantly is rheumatic heart disease [10]. In the cardiac follow up clinic of Jimma Specialized Hospital, rheumatic heart disease was the leading cause of cardiac illness (32.8% of all cardiac cases) followed by hypertensive heart disease (24.2%), cardiomyopathy(20.2%), arrhythmia in 13.5% and Cor-pulmonale accounting for 3.8% of all cardiac cases[11]. A study conducted in six main Referral Hospitals of Ethiopia also showed that RHD is the common cause of cardiovascular disease followed by HHD and cardiomyopathy [12]. However, there is an increasing trend of hypertensive heart disease in the subsequent periods taking the place of valvular diseases.

A recent study conducted in Northern Ethiopia focusing on the trend of cardiovarcular diseases in outpatients showed that Hypertensive heart disease was the predominant etiologic diagnosis of cardiovascular disease followed by rheumatic heart disease [13]. In this study, hypertensive heart disease had surpassed rheumatic heart disease as the leading heart disease. This might be due to the increasing prevalence of hypertension, as supported by the high proportion of hypertension of approximately two thirds (62.3%) of CVDs found in this study [13]. In another African study the main causes of heart failure were hypertension (21.3%), rheumatic heart diseases in 20.1% (with the commonest form being mitral regurgitation(78%)), cardiomyopathy in 16.8% (with dilated crdiomyopathy being the commonest form of ideopathic cardiomyopathy (67.7%)), coronary artery disease(10%), and congenital heart disease (9.8%)[14]. Studies showed that much of the population risk of CVD is attributable to modifiable traditional risk factors, including smoking, hypertension, diabetes, and obesity, lack of physical activity, raised blood lipids and psychosocial factors [6, 15]. These risk factors account for 61% of CVD deaths globally and alleviating exposure to these risk factors would improve global life expectancy by almost 5 years [16]. Data availability on CVDs and their risk factors remains low in Ethiopia, making accurate estimation of burden in terms of the morbidity and mortality of CVDs extremely difficult. This study will therefore provide relevant data regarding the magnitude as well as spectrum of cardiovascular admission and its outcome in st. Paul specialized hospital, Addis Ababa.

## Methods

### Study setting

The study was conducted at St Paul Millennium Medical College (SPHMMC) which is one of the tertiary governmental Teaching Hospitals in Addis Ababa, the Capital of Ethiopia and the seat of African Union, and the second largest public health Hospital in Ethiopia. The hospital receives follow-up patients, emergency patients and referrals from other Hospitals in the country and health centers in the catchment area. The study was conducted in the medical department of the Hospital on all cardiovascular admissions among the medical admissions from 1st of January of 2020 to January 1of 2021. The hospital has a bed capacity of 350, of which 45 beds are allocated to medical wards and 14 beds for ICU. Among others, It has 8 medical regular outpatient departments(OPDs) for follow up patients working the whole weekdays and big adult medical emergency OPD providing care 7/24.

### Study design

An institutional based, descriptive, retrospective, cross-sectional study was conducted at St Paul Teaching Hospital on all cardiovascular admissions among the medical admissions in the medical ward and ICU from 1st of January 2020 to 1st of January 2021. Admission and discharge diagnosis was used from the registry to retrieve the chart of the patient for detailed review of demographic data, major investigations, co-morbidities, underlying background risk factors, and previous treatment history and follow up.

### Study population

The study included all eligible cardiovascular admissions among all the medical patients admitted at the study hospital from 1^st^ of January 2020 to January of 2021. A total of 353 patients with cardiovascular diagnosis among the annual medical admissions of 1165 patients were evaluated during the specified study period. The exclusion criteria were patients whose medical records were lost, incomplete data, or patients died on arrival before adequate diagnosis was made.

### Data Variables for the study

Patient information was obtained from the HMIS registry as well as medical records to include dates of admission and discharge or death or referral/transfer or left against medical advice (LAMA), demographic data, diagnosis (admission diagnosis, final diagnosis including co-morbidities), and total duration of hospital stay. Outcome variables were the status of the patient at discharge from the hospital such as discharged with improvement, referral to other hospitals, LAMA-left against medical advice, and death in the hospital.

Cardiovascular diseases are diseases that affect the heart and blood vessels. The main blocks of WHO International Classification of Diseases(ICD)-10th version for Mortality and Morbidity Statistics (MMS) (Version: 04 / 2019) was utilized to sort out the final diagnoses [18]. The unit of analysis was the hospital discharge and/or admission, not the patient and therefore a patients discharged more than once in a year were counted each time as a separate “discharge” from the hospital. In situations with more than one cardiovascular diagnosis in the same case, the different disease conditions were counted separately.

### Operational definition

**Cardiovascular diseases** comprise various diseases affecting the heart and blood vessels, resulting in different comorbidities and life-threatening complications.

**Cardiovascular admissions** were patients among all medical admissions during the stated period having one or more diagnosis related to the cardiovascular diseases as documented in the HMIS. Some of such patients had cardiovascular illnesses as a sole diagnosis and others had related or unrelated medical co-morbidities.

**Hypertensive heart disease (HHD**) was diagnosed in patients with hypertension presenting with symptoms and signs of heart failure, with or without left ventricular (LV) hypertrophy and left atrial enlargement on two-dimensional echocardiographic evidence of LV diastolic dysfunction in the absence of significant valvular heart disease or regional wall motion abnormality[12]. For those who have no documented 2-dimentional echocardiogrpahy, ECG evidence of left ventricular hypertrophy (LVH) was used.

**Dilated cardiomyopathy** was diagnosed in patients with marked LV dilatation and dysfunction in the absence of significant valvular, structural, or congenital heart disease or arterial hypertension[12].

**Ischaemic heart disease (IHD)** included three entities: 1. Angina pectoris which is short lived, relieved with termination of the provoking factor or rest and had no typical ECG features of infarction, 2. Acute myocardial infarction which was defined by the presence of elevated cardiac biomarker together with acute onset chest pain, and/or typical ECG changes, 3. Prior myocardial infarction including patients who present with or without heart failure in whom echocardiography detected regional wall motion abnormality in the absence of a history of acute coronary syndrome[12]. Pathologic Q waves on ECG were used to predict the possibility of ischaemia as a cause of the regional wall motion abnormality in a dilated left ventricle with reduced ejection fraction.

**Pulmonary heart disease (PHD)** referring to right heart failure also known as cor pulmonale due to altered structure and/or function of the right ventricle evidenced by 2-dimensional echocardiography. This Heart disease could be due to pulmonary hypertension secondary to disease of the lungs (Eg. COPD) or its blood vessels (Eg. pulmonary thrombo embolism). PHD also included primary pulmonary diseases.

#### Blood pressure control

Controlled BP when the BP < 140/90 mmHg in hypertensive patients of all ages [41]. And Uncontrolled BP when the BP ≥ 140/90 mmHg in hypertensive diabetic patients of all ages [41]

#### Blood Sugar control

Good glycemic control when the mean Fasting blood sugar ≤ 130 mg/dL and /or HbA1C <7% and Poor glycemic control when mean Fasting blood sugar >130 mg/dL and/or HbA1C>7%[42]

### Data collection and instrument

In order to ensure the accuracy, completeness, and comparability of data, four final year medical residents were trained to complete data collection format. Data collection was made by the pretested structured check lists to document all the pertinent profiles of the study subjects.

### Data Processing and Statistical Analysis

After checking for completeness, data was coded, entered, and analyzed using SPSS version 20 software. Descriptive statistics was used to calculate rates. Chi-squares was used to estimate the associations between selected predictor variables. A p-value < 0.05 was taken as statistically significant.

### Ethical Consideration

Ethical clearance was obtained from institutional review board of St. Paul Teaching Hospital. Since it was a retrospective study, written consent was not obtained. Anonymity of the patient profile was upheld. Ethics and research committee approval from the Institutional Review Board (IRB) was obtained.

## Results

### Socio-demographic characteristics of the study population

From the annual medical admissions of 1,165 patients to st. Paul Hospital Mellenium Medical College from January 1 2010 up to January 1 of 2021, the total cardiovascular admission constituted 34%(395). However, the missing charts of 11%(40) were excluded making the cardiovascular admissions of 30.3%(353) utilized for further scrutiny. Majority of the study subjects were females (60%(213)) and about half of the patients were urban residents of Addis Ababa. From the 353 cardiovascular admissions, 40.5%(143) were from Oromia regional state and the rest of the patients were referred from other six regional states of the country as well as Dire Dawa. Therefore, Addis Ababa together with Oromia regional state constituted 90% of total cardiovascular admissions of St. Paul Hospital Millennium Medical College during the study period (Table. 1). The minimum age among the cardiovascular admissions was 15years and the maximum age was 86years with the mean age of 48.9years, and the median age was 50years. The overall cardiovascular illness increased almost steeply with increasing age(p=0.001) (fig. 1). However, further disease stratification showed that Rheumatic valvular heart diseases(CRVHD) and vascular diseases including DVT, PTE and CVT were more common in the younger age and proportionally decreased as age advanced(p=0.001) (Fig 2).

**Table 1.**
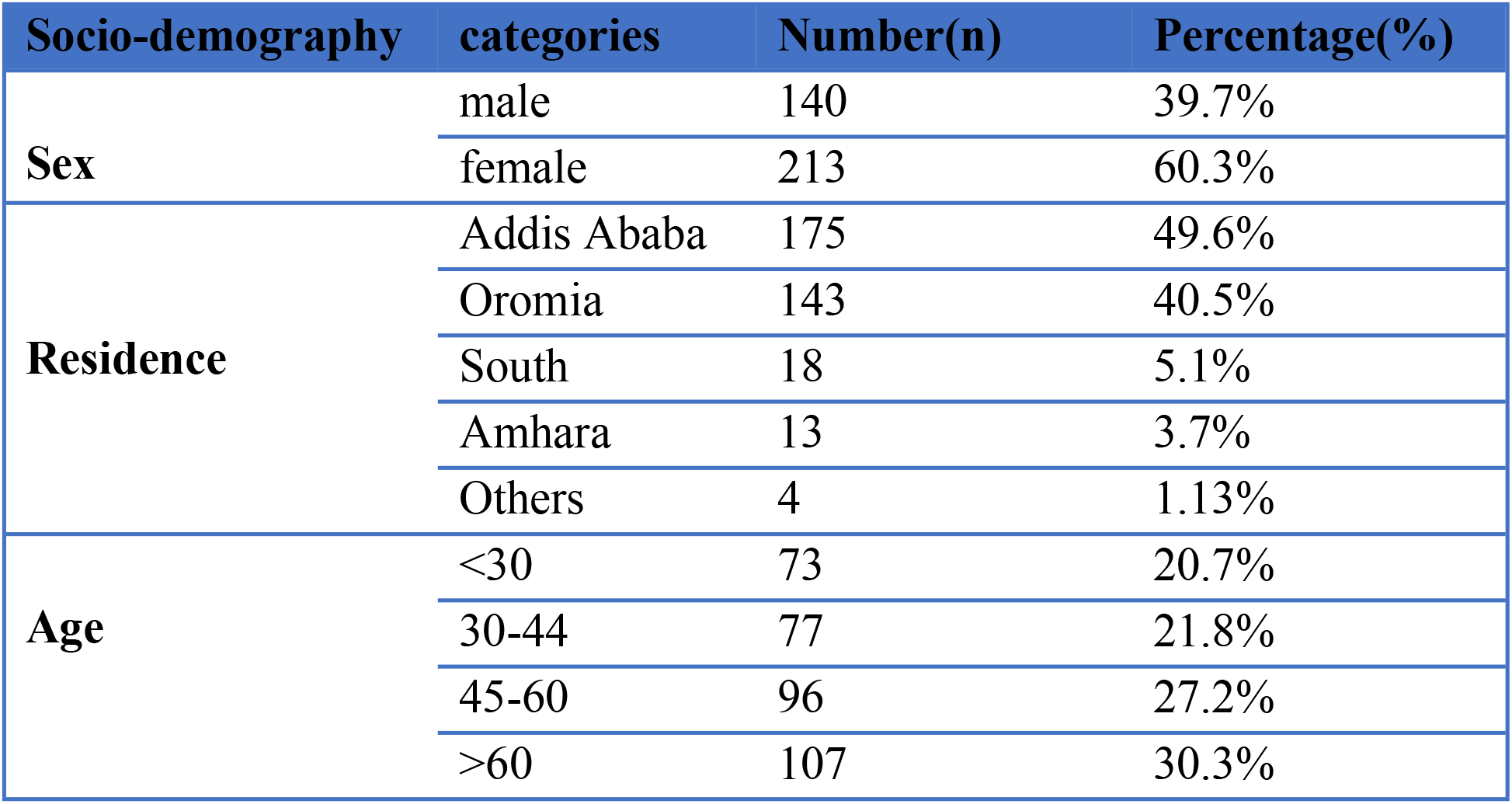
Socio-demographic characteristics of cardiovascular admissions in St Paul Hospital Millennium Medical College

| Socio-demography | categories | Number(n) | Percentage(%) |
| --- | --- | --- | --- |
| Sex | male | 140 | 39.7% |
|  | female | 213 | 60.3% |
| Residence | Addis Ababa | 175 | 49.6% |
|  | Oromia | 143 | 40.5% |
|  | South | 18 | 5.1% |
|  | Amhara | 13 | 3.7% |
|  | Others | 4 | 1.13% |
| Age | <30 | 73 | 20.7% |
|  | 30-44 | 77 | 21.8% |
|  | 45-60 | 96 | 27.2% |
|  | >60 | 107 | 30.3% |

**Fig 1.**
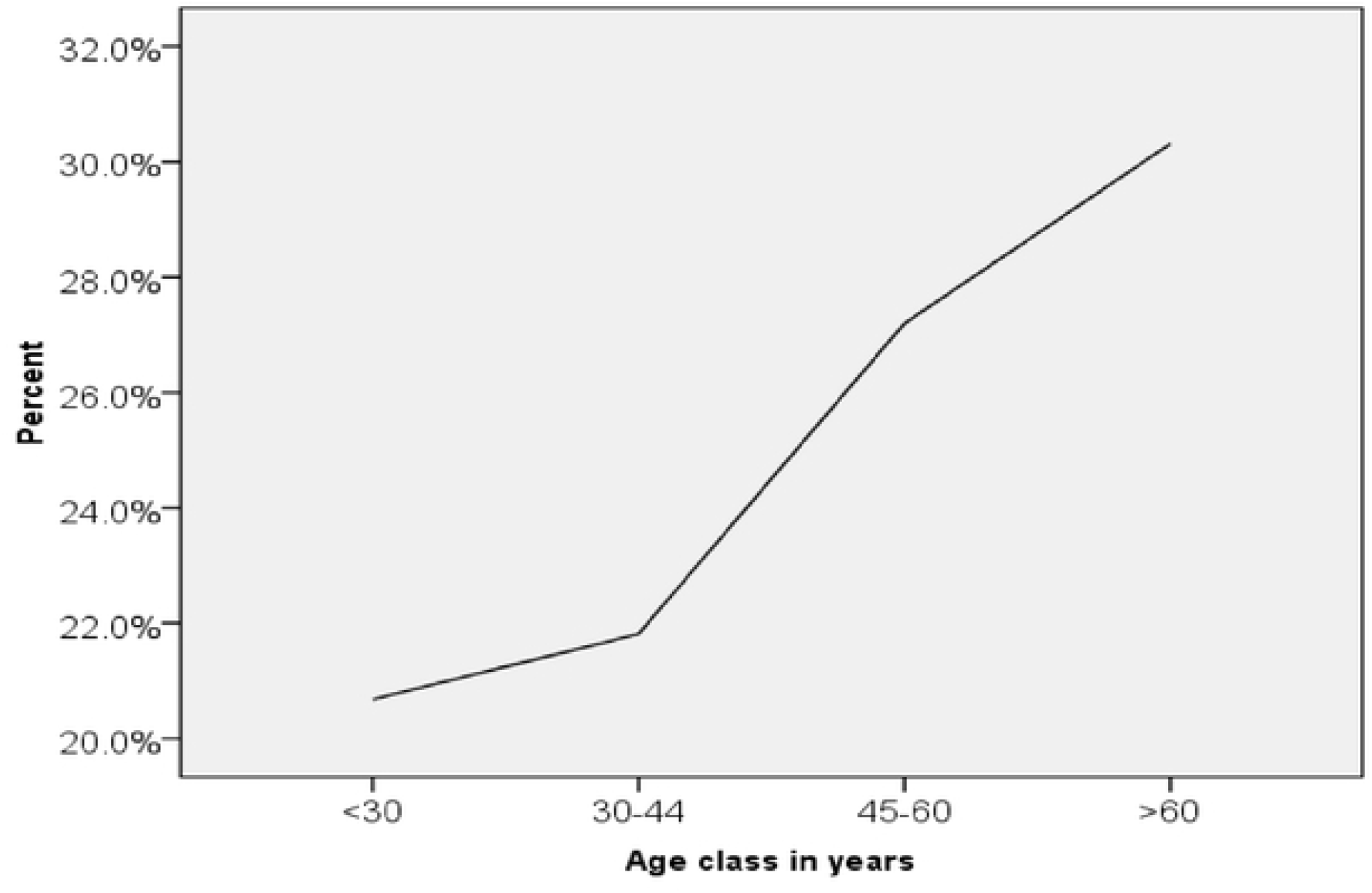
The relationship of the overall cardiovascular admissions with age. The x-axis shows the age class and Y-axis shows the percentage of CVD burden in each age group.

**Fig 2.**
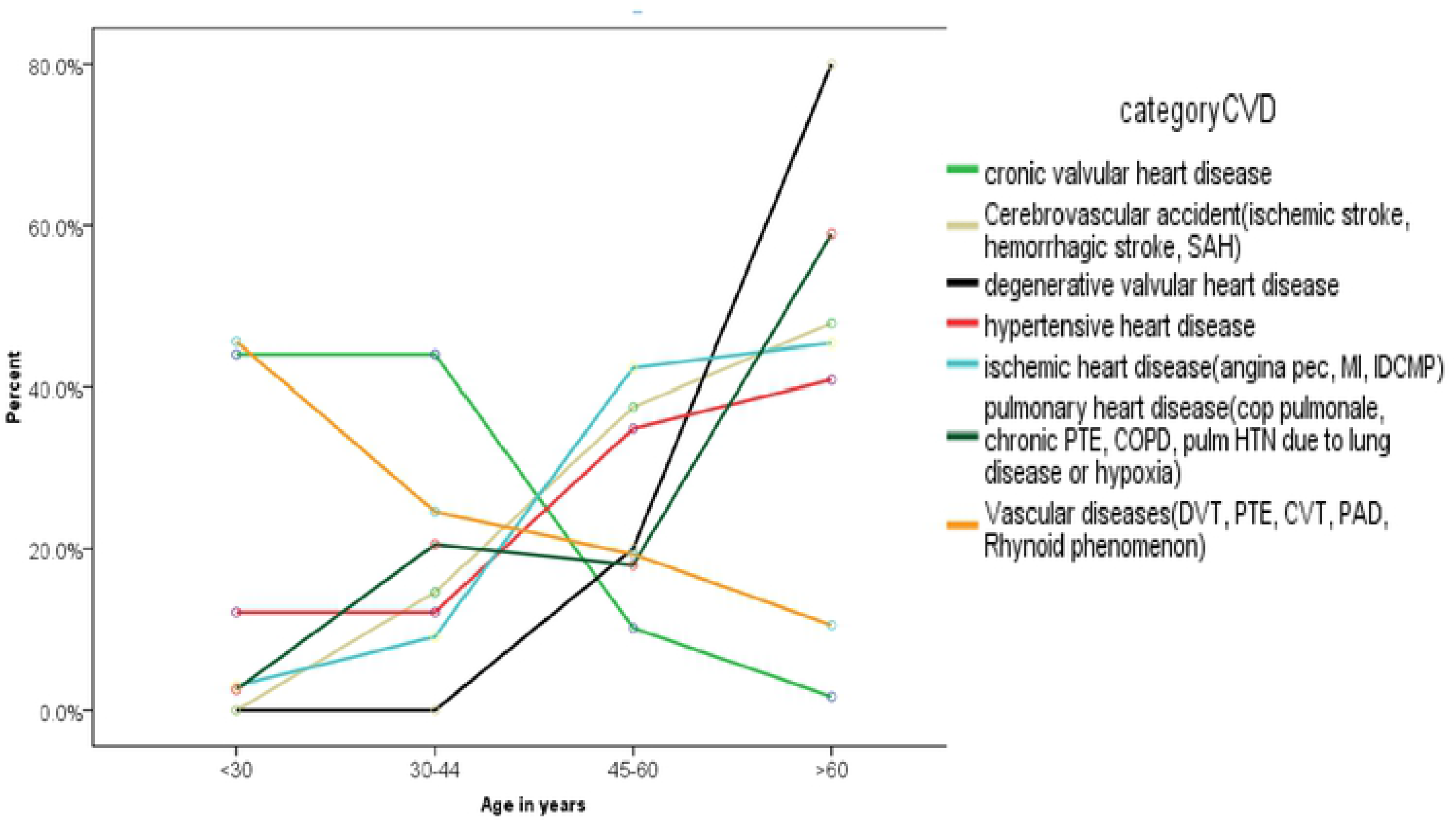
The associations of major cardiovascular diseases with increasing age. p<0.001.

**Fig 3.**
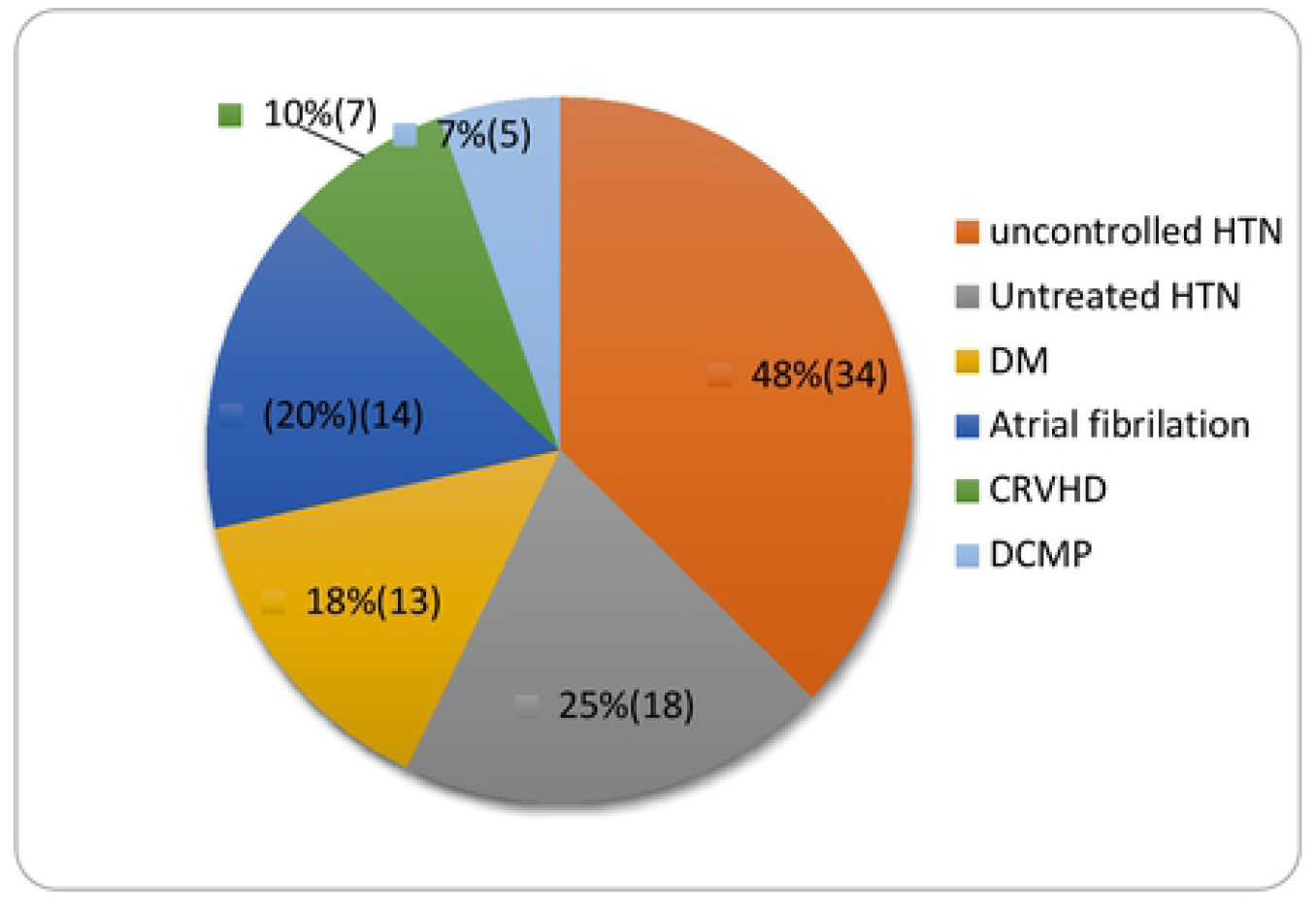
Major risk factors of stroke among admitted patients in St Paul Hospital Millennium Medical College.

**Fig 4.**
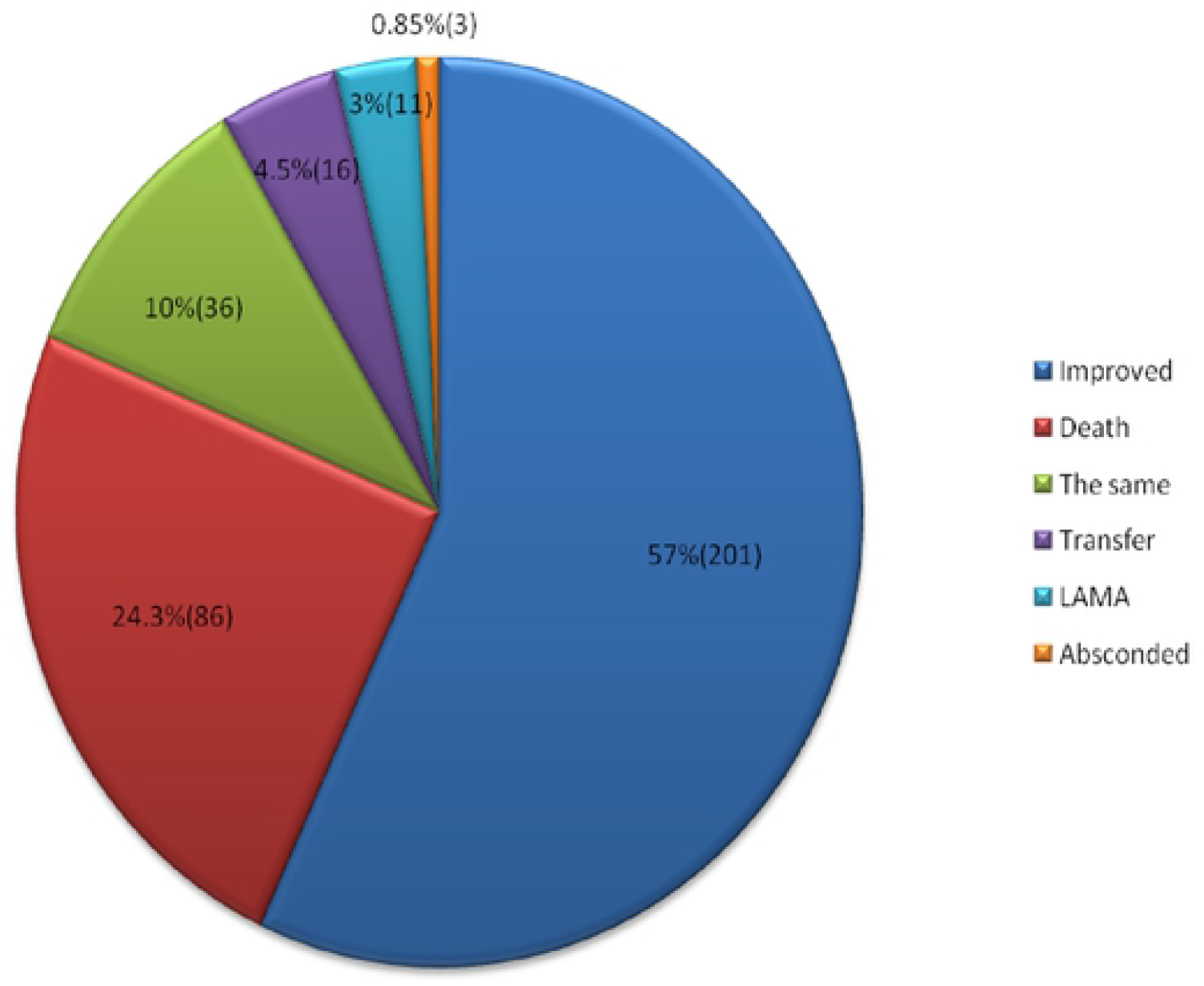
The outcomes of cardiovascular admission in St Paul Hospital Millennium Medical College.

### Spectrum of cardiovascular diseases and associated risk factors

The spectrum of major cardiovascular diseases was scrutinized according to the ICD-10 classification of diseases and most of the patients had two or more cardiovascular diagnosis. From all cardiovascular admissions, about 60%(212) of patients had advanced congestive heart failure (NYHA class III and class IV) of diverse causes. The advanced congestive heart failure (CHF) had multiple underlying etiologies, and also multiple precipitating factors including hypertension(35%(74)), lung infection (34%(72)), drug discontinuation (25.5%(54)), and arrhythmia (23.6%(50)) predominantly being atrial fibrillation with fast ventricular response(Afib with FVR).

Hypertensive heart disease (HHD) constituted for 41.4%(146) of the total admission diagnosis as well as the leading cause of heart failure and its prevalence increased with increasing age (p<0.001). HHD was more predominant in urban residents of Addis Ababa than those who came from out of Addis Ababa (63%(92) Vs 37%(54), p=0.001). Valvular heart diseases (both rheumatic and degenerative valvular heart diseases) accounted for about 34.5% of all advanced cardiac failure cases and 20.5% (73) of all the cardiovascular admissions as shown in the Table 2 below. Chronic rheumatic valvular heart diseases(CRVHD) was more dominant in patients coming out of Addis Ababa compared to residents of Addis Ababa(82.8%(53) Vs 17.1%(11)(p=0.001). Echocardiography proven isolated MS and MS with MR constituted 26%(19) and 27.5%(20) of all valvular heart diseases, respectively. There were 20.5% of isolated MR cases and 13.5% of mixed MS, MR, and AR of all valvular heart diseases, with the rest comprising 12%. The proportion of degenerative valvular heart diseases increased with advancing ages (p=0.01) whereas the proportion of rheumatic heart diseases decreased as age increasing (p=0.01).

**Table 2.** Spectrum of major cardiovascular Diseases.

| **CVD spectrum** | **Frequency(n)** | **Percentage(%)** |
| --- | --- | --- |
| **CHF** | 212 | 60 |
| **HHD** | 146 | 41.36 |
| **CRVHD** | 64 | 18.1 |
| **IHD** | 43 | 12.2 |
| **PHD** | 40 | 11.3 |
| **DCMP** | 34 | 9.6 |
| **DVHD** | 9 | 2.5 |
| **Vascular DiseasesΔ** | 85 | 24 |
| **CVA** | 70 | 20 |
| **ARRYTH** | 68 | 19.2 |
| **SBE** | 10 | 2.8 |
| **OTHERS\*** | 31 | 8.8 |

\*Thyocardiatic disease, Intra-cavitary masses, High output Heart failure, ΔPAD, DVT, PE, CVT

Ischemic heart disease (IHD) accounted for 12.2%(43) of all CVD admissions and the prevalence has significantly increased with age and males had significantly higher proportion than females(p=0.001) (Table 3). IHD was more common in residents of Addis Ababa than those who came from out of Addis Ababa but the difference was not significant (p=0.2). IHD is associated with hypertension and diabetes (p=0.001). From all IHD patients, 90.24% had hypertension with 13.5% had good hypertension control, 59.5% had poor hypertension control, and 27% were newly diagnosed for hypertension or not on treatment. Similarly, 41.5% of IHD patients had T2DM with 41.5% had good glycemic control, 35.3% had poor glycemic control and 17.6% were newly diagnosed for T2DM. Over 42% of pulmonary heart disease (PHD) was attributed to COPD related causes and the remaining was due to chronic PTE and secondary to post-tuberculosis fibrosis. PHD was more common in patients who came from Addis Ababa then those who came from out of Addis Ababa but it was not significant (p=0.053). Females constituted over two-third of dilated cardiomyopathy and peripartal cardiomyopathy accounted for 26.5%(p=0.01).

Vascular diseases (VasD) were the most common cause of cardiovascular diseases next to CHF and HHD. It accounted for about a quarter of cardiovascular diagnosis and females constituted about two-third of the cases. DVT was the leading vascular cause accounting for 44%(37) followed by PTE of about 30%(25). Peripheral arterial disease (PAD) and cerebral venous thrombosis(CVT) accounted for 15.5%(13) and 9.5%(8) of vascular disease admission, respectively. Pregnancy related conditions including caesarian section and puerperium (30.6%(26)), major surgery and prolonged immobilization for medical illnesses(28.2%(24)) as well as active cancers(11.7%(10)) were the major risk contributors of venous thrombo-embolism(VTE). Residence of the patients did not have effect on the distribution of vascular diseases (P=0.1). Hypertension and diabetes were significantly associated with PAD(p=0.001). Hypertension was documented as a risk factor in about 85% of patients with PAD followed by Type-2 diabetes mellitus and chronic kidney disease, each of which were implicated as a risk factor in 25% of cases of PAD. The cumulative vascular diseases significantly decrease as the age of the patients increased(p=0.01)(Fig. 2).

Cerebrovascular accident (Ischemic stroke and hemorrhagic stroke) accounted for about 20%(71) of total annual cardiovascular admission with hemorrhagic stroke constituted about 17% and ischemic stroke for the remaining 83% of the total stroke patients. Of all ischemic stroke, cardioembolic stroke amounted for 32.2%. From all the atherothrombotic ischemic stroke groups, 7.5% had hemorrhagic transformation during the course of disease progression. Stroke was predominant in residents of Addis Ababa than those who came out of Addis Ababa albeit the difference was not statistically significant (p=0.07) but it increased with increasing age(p=0.001). There was no variation of stroke distribution between males and females (male=45% and female=55%, p=0.09).

All patients with hemorrhagic stroke had hypertension and 75% of them had long standing uncontrolled hypertension whereas the remaining 25% had untreated hypertension; either newly diagnosed hypertension or known hypertension on life style modification self-deferring the pharmacologic therapy. Similarly, 97.5% of atherothrombotic ischemic stroke patients had hypertension with 61.5% of them had uncontrolled hypertension whereas about 38% had untreated hypertension. About one-third of atherothrombotic ischemic stroke(non-cardio-embolic) patients had type-2 diabetes. Atrial fibrillation accounted for 73.7% of cardioembolic stroke admissions followed by CRVHD (36.8%) and DCMP (26.3%), respectively. About 71.5% of CRVHD patients and 60% of DCMP patients had atrial fibrillation.

### Hypertension and Diabetes control among the cardiovascular admissions

Of all the 353 annual cardiovascular admissions, 47.6%(168) patients had history of hypertension. Hypertension is significantly prevalent in patients from Addis Ababa than patients who come out of Addis Ababa ((63.7%(107) Vs 36.3%(61), p=0.001)). The minimum duration of hypertension ranged from newly diagnosed hypertension up to the maximum years of hypertension history of 30 years. Only 20% of patients reported to have good hypertension control (BP<140/90mmHg) with adherence to the medications and have frequent follow up whereas 46.6% of patients had poor hypertension control (BP≥140/90mmHg) who were either not adherent to or discontinued their pharmacologic therapy by themselves and came with end organ damage. About 35% of hypertensive patients were not on any sort of therapy or diagnosed to have hypertension after hospital presentation for their current respective complaints. The control of hypertension in patients from Addis Ababa was significantly better than control in patients coming from Addis Ababa (p=0.001). The proportion of diabetes mellitus(DM) among the cardiovascular admissions were 17.6%(62) and significantly prevalent in patients from Addis Ababa than patients who came from out of Addis Abbaba (74.1%(46) Vs 25.8%(16), p=0.001). The duration of DM history ranged from newly diagnosed T2DM after hospital admission up to 25 years. About 40% of DM patients had ‘‘good’’ DM control with FBS≤ 130mg/dl and/or A1C<7% where as 46.8% had ‘‘poor’’ DM control (FBS>130mg/dl and/or A1C>7%) having erratic adherence to the medication, the initial single drug as well as the dose never changed and had no follow up. Most of the patients had no documented HbA1c. The control of diabetes in patients from Addis Ababa was significantly better than control in patients coming from Addis Ababa (p=0.001).

### Outcomes of cardiovascular disease admissions

Among patients with CVD admissions, the minimum hospital stay was one day and the maximum hospital stay was 80 days with the average hospital stay was 12days. About 57%(201) of cardiovascular admission patients were discharged from the hospital with improvement but in hospital mortality rate among the cardiovascular admission was 24.3%(86). The immediate causes of death were sepsis with septic shock(25.6%), fatal arrhythmia (19.8%), brain herniation (15%), massive PTE (14%), and cardiogenic shock (11.6%) (Table 4).

**Table 4.**
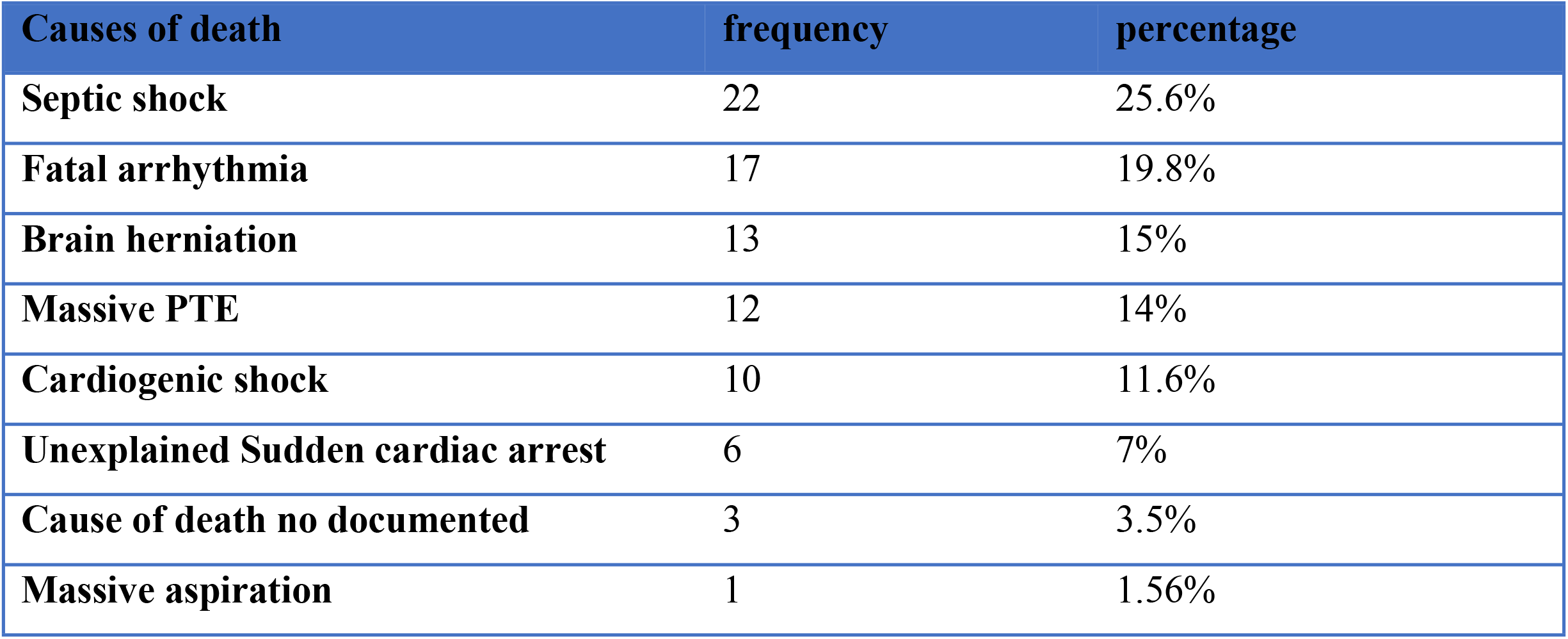
Major causes of death of cardiovascular admissions.

## Discussion

The epidemic of cardiovascular disease (CVD) is a global phenomenon, and the magnitude of its burden is alarmingly increasing in the developing world including Ethiopia. The cardiovascular disease remains the leading cause of death in the world and approximately 80% of all CVD-related deaths occur in low and middle income countries [6]. In the current study which involved the annual medical admissions of 1,165 patients at st. Paul Hospital Millennium Medical College, the total cardiovascular admission constituted 30.3%. The present burden of cardiovascular cause of medical admission was in agreement with the previous study conducted in the same hospital in which the CVD prevalence was found to be 32% among the total medical admission [17]. However, the previous study considered hypertension as a separate exclusive CVD diagnosis but in this study hypertension was considered as a CVD risk factor and hypertensive heart disease was an exclusive diagnosis as per the International Disease Classification(ICD-10)[18]. The present finding is also similar with the cardiovascular admission of 31% from all medical admission in the Nigerian Teaching hospital[19]. However, the current rate of CVD admission is much greater than 17.5% of CVD admission in Asella Referal Hospital [20]. This higher rate in our study might be due to large proportion of urban residency dominated by cardiovascular risk factors including hypertension and diabetes unlike the former study catchment area where infectious cause of admission accounted for close to 50% of hospital admission[20].

Most of the patients (60%) were admitted with the clinical diagnosis of advanced congestive heart failure (CHF) (NYHA class III and IV) having multiple causes and with chief complaint of worsening of shortness of breath, orthopnea, PND and body swelling. This high rate of CHF in the current study is in agreement with other study [17, 21]. Surprisingly, over a quarter of CHF patients had associated history of drug discontinuation as a precipitating factor eventually worsening their heart failure condition. Therefore, healthcare providers should also equally focus on health education including adherence to the medical managements in addition to prescribing the pharmacologic therapy based on the appropriate diagnosis.

Hypertensive heart disease (HHD) was the second predominant admission diagnosis constituting for 41% of overall cardiovascular admission and also the leading cause of heart failure followed by rheumatic valvular heart disease(RVHD) which constituted for about 18% of all CVD. In most previous studies involving both inpatient medical patients and outpatient follow up clinics, VHD was considered to be the leading cause of CVD in general and cardiac diseases in particular [11, 13, 17, 20, 21, 22]. This increasing trend of HHD shown in the current study is consistent with the study done in Gondar Hospital [13]. This might be due to the increasing prevalence of hypertension, as supported by the high proportion (about 48%) of hypertension among CVD patients found in our study. HHD in the current study was predominant in urban residents than the rural dwellers (p=0.01). It may be an indication of increasing urbanization and westernization of diet in the present metropolitan area coupled with better health care availability in the current urban area disfavoring the risk factors for rheumatic valvular heart disease (RVHD). RVHD in the current study is dominant in patients coming from rural area than the residents of Addis Ababa(p=0.001). Rural predominance of RVHD is consistent with previous studies [11, 12, 13]. This could partly be justified by the early treatment of respiratory tract infection and better access to health care in urban residents. RVHD in the present study was also predominant in females (p=0.001) and it is in agreement with previous studies[11, 12]. Ischemic heart disease (IHD) and dilated cardiomyopathy(DCMP) were other causes of cardiac diseases constituting for 12.2% and 9.6% of annual CVD admissions, respectively and the burden increased with increasing age(p=0.01). These findings are in agreement with other national studies [12, 13, 21]. The ischemic heart disease has increased when the current rate of 12.2% (12,200 per 1000,000) among the CVD patients and the 2014 IHD rate of 7,400 per 100,000[10] is compared with reports of 640 per 100,000 in 1993 [23] and 88 per 100,000 in 1988 [40]. Hypertension and Diabetes mellitus were highly associated with the development of IHD (p=0.001) and is in agreement with similar study[10].

Cerebrovascular accident (CVA) including both ischemic and hemorrhagic stroke accounted for 20% of annual CVD admissions with ischemic stroke dominated by 83% whereas hemorrhagic stroke taking the remaining 17%. This finding is consistent with the long established global data of 85% ischemic stroke versus 15% hemorrharic stroke and some Ethiopian data where Ischemic stroke is the dominant sub-type[10, 24, 25]. A study done by abdissa *et al* [10] at Tikur Anbessa hospital showed that Hemorrhagic stroke accounted for about 37.5% and ischemic stroke for 55% of total stroke patients. However, the current finding is not in agreement with some Ethiopian data that reported the hemorrhagic stroke markedly outnumbered the ischemic stroke [26, 27, 28]. The markedly exaggerated hemorrhagic stroke compared to ischemic stroke which was reported in a number of studies in Ethiopia mandates further scrutiny. Since most of the stroke studies conducted so far were in the admitted patients in the hospitals, the findings may not be representative of the actual relative proportions of the sub-types of stoke as it is routine phenomenon that most patients with mild or moderate ischemic stroke do not necessarily come to hospital attention or might have been early discharged from ER with no admission once the acute phase is over or due to their late presentation after the acute phase [29].

In this study, hypertension was present in 74% of patients with stroke. All hemorrhagic stroke patients had hypertension and 75% of them had long standing uncontrolled hypertension whereas the remaining 25% had untreated hypertension. This finding is in agreement with similar studies[24, 26,30]. Furthermore, 97.5% of atherothrombotic(non-cardioembolic) ischemic stroke patients had hypertension as a major risk factor where 61% of them had uncontrolled hypertension and 38% of them had newly diagnosed or untreated hypertension. The find is also in conformity to other studies [24, 26, 30]. Therefore, poor medical follow up and poor pharmacologic adherence against CVD risk factors including hypertension, makes it imperative to revisit our management practices and protocols, so as to establish a consolidated approach involving the relevant stakeholders. For countries of poor economy including ours, prevention is much cheaper and profitable than the complex and costly managements of the subsequent CVD diseases and their complications.

Hypertension, in the current study, was found to be one of the dominant risk factor for cardiovascular diseases where about 48% of CVD patients had hypertension. However, this finding is higher than some of the previous reports of hypertension burden among the CVD patients in Ethiopia[10, 11, 12, 17, 31] but much lower than the 62% as shown in the study conducted in Gondar Referral Hospital[13]. Our finding is also consistent with the overall increasing trends of atherosclerotic CVD burden in the recent decades with hypertension being a significant risk factor [13, 32]. The worrisome finding was that only 20% of our patients with longstanding hypertension had good hypertension control whereas about 47% of patients had poor hypertension control who were either not adherent to or discontinued their antihypertensive therapies by themselves. Furthermore, 35.2% of hypertensive patients were not on any sort of anti-hypertensive therapy or diagnosed to have hypertension after hospital presentation for their current respective complaints. This poor hypertension control is consistent with other studies [33, 34] and warrants timely intervention. Similarly, from all CVD patients with long standing diabetes mellitus, only 40.3% had good glycemic control where as 46.8% had poor glycemic control and poor follow up. About 13% of diabetes was discovered as newly diagnosed type-2 diabetes after they were admitted with the current cardiovascular diseases. These findings are consistent with other studies in Ethiopia [35, 36] and much better than the 20% of good glycemic control at Tikur Anbessa Specialized Hospital [42].

Vascular diseases including deep venous thrombosis (DVT), pulmonary thrombosis (PTE), peripheral arterial disease(PAD), and cerebral venous thrombosis (CVT) were very rampant in the present study area and all together constituted about 24% of all the annual CVD admissions. DVT was the leading vascular cause forming a 44%, followed by PTE of 30%, PAD of 15.5% and CVT for 9.5% of vascular disease admission, respectively. The current burden is much higher than the previous cardiovascular spectrum studies [10, 12, 13] which involved the out patients, unlike the admitted patients who have higher risk factors[37, 38]. Pregnancy and its related conditions such as puerperium, and cesarean section (30.6%), major surgery and prolonged immobilization for medical illnesses (28.2%), together with active cancers(11.7%) were the major risk contributors of VTE. The higher burden of DVT; younger age and female predominance; major risks including pregnancy and puerperium, prolonged immobility and malignancy; documented in the current study are consistent with reports from prospective studies in Addis Ababa Hospitals [37, 38]. The high burden of VTE including massive PTE as one of the major immediate cause of death (Table 4) may be partly due to lack of comprehensive guideline based venous thromboembolism(VTE) prophylaxis consistent with the degree of risk factors. The tradition of prolonged bed rest during puerperiuma might have also exacerbated the existing procoagulant risks. Hypertension was documented as a risk factor in about 85% of patients with PAD followed by Type-2 diabetes mellitus implicated in 25% of cases of PAD. This is consistent with the association of uncontrolled hypertension and diabetes with atherosclerotic CVD diseases including PAD[31]. Therefore, such findings mandate the consideration of comprehensive approach ranging from simple and cost effective but targeted health education to break the immobility segment of Virchow’s triad particularly during puerperium in which women in most part of Ethiopia are customarily advocated to rest for protracted days or weeks after delivery by potentially increasing the risk of VTE, through adequate risk assessment for thromboprophylaxis in hospitalized patients for medical or surgical admissions, and adequate anticoagulation to prevent recurrence of VTE.

The mean duration of total hospital stay in the current study was 12days with the minimum hospital stay was one day and the maximum hospital stay was 80 days. This result is comparable to the average of 12.33days of hospital stay for medical admission at st. Paul hospital [17] stroke admissions of 11.14days [27] but much less than the 14.4days of total hospital stay at Asella referral hospital[20]. There were 57% of CVD patients discharged with improvement and in hospital mortality rate was 24.3%. These finding are comparable to the previous st. paul hospital reports of in hospital death rate of 24.2% and discharge rate with improvement of 66% among the medical admissions. Despite the current mortality rate is lower than the 25% documented among the cardiovascular admissions in Addis Ababa [21] and the 30% mortality rate among stroke admissions in st. Paul Teaching hospital [27], it is much higher than the 12.2% among cardiovascular admissions in Nigeria [19]. Sepsis with septic shock was the leading immediate cause of death attributable for about a quarter of deaths among CVD admissions, followed by fatal arrhythmia, brain herniation, massive PTE and cardiogenic shock in 20%, 15% and 14%, respectively. Infectious disease was also the leading cause of mortality among medical admissions studied previously [17, 20]. Therefore, comprehensive work is ahead both at institution level and national level to lower the CVD risks, disease progression, fatality rate and the duration of hospital stay.

In conclusion, cardiovascular diseases are highly prevalent in the study area causing significant morbidity and mortality. Most cardiovascular diseases have preceding unaddressed but easily modifiable risk factors, controlling which could have prevented or delayed the progression of the disease. Therefore, concerted effort is required from all stake-holders in terms of risk reduction, follow-up optimization to prevent or delay cardiovascular disease development, disease progression and subsequent complications.

## Data Availability

All relevant data are within the manuscript and its Supporting Information files.

## Author’s contribution

Dr. Mekoya: Conceptualization, methodology, data collection, write up

Dr. Henok: Methodology, write up

